# Prevalence of somatosensory modulation of the cervical spine and temporomandibular joint in subjects with tinnitus: a systematic review

**DOI:** 10.1101/2023.08.26.23294605

**Authors:** Marco Segreto, Andrea Giusti, Paolo Bizzarri

## Abstract

**Background:** Tinnitus is defined as the perception of a sound without a corresponding external acoustic stimulus and is considered a symptom rather than a disease. In some individuals, it can be evoked or modulated by input from the somatosensory, somatomotor, and visual-motor systems. This has led to the introduction of the term: ‘‘somatosensory modulation of tinnitus’’. In these cases, the psychoacoustic attributes of tinnitus (loudness and pitch) can change as a result of external stimuli such as strong contractions of the muscles of the head, neck, limbs, orofacial movements, and eye movements. The temporomandibular joint and cervical spine, along with the head, appear to be the musculoskeletal anatomical regions most commonly underlying somatic tinnitus. This review aims to evaluate the prevalence of somatosensory modulation of the cervical spine and temporomandibular joint in subjects with tinnitus.

**Methods:** The databases investigated for the review were: Embase, Pubmed, Scopus, and Web of Science. Observational studies investigating the prevalence of somatosensory modulation of the cervical spine and temporomandibular joint in subjects with tinnitus were included. No time limit was entered into the search and no age or language restrictions were applied.

**Results:** 14 studies met the eligibility criteria on which the review was based. In 5 studies, the prevalence of tinnitus modulation was reported with one or more maneuvers involving TMJ, in 3 studies with one or more maneuvers involving the cervical spine and 7 studies reported the prevalence of modulation following maneuvers for both somatic regions.

**Conclusion:** The present study confirmed that tinnitus perception and intensity can be somatically modulated in a subpopulation of individuals. The cervical spine and the ATM play a key role in the modulation of tinnitus and, based on the data collected on individual districts, the ATM appears to be the most frequently involved somatic region.

## 1. INTRODUCTION

Tinnitus is defined as the perception of a sound without a corresponding external acoustic stimulus [1,2] and is considered a symptom rather than a disease [3].

Tinnitus has a prevalence of 11.9 -30.3% in the adult population [4], but only 1-3% report it as a condition that worsens quality of life [5].

Social factors such as low income, poor education, or exposure to noise during occupational and recreational activities can influence the prevalence of tinnitus [6].

Tinnitus is typically associated with hearing loss (which can be experienced in up to 90% of cases), use of ototoxic drugs, infections, and other medical conditions that can affect hearing function by triggering cochlear damage, resulting in neural changes in the central auditory system [6,7]. In such patients, tinnitus is referred to as ‘‘otic tinnitus’’ [2].

However, tinnitus can be evoked or modulated by input from the somatosensory, somatomotor, and visual-motor systems in some individuals [8,9]. This has led to the introduction of the term: ‘‘somatosensory modulation of tinnitus’’ [8]. In these cases, the psychoacoustic attributes of tinnitus (loudness and tone) can change, even if often only temporarily, following external stimuli such as strong contractions of the muscles of the head, neck, and limbs [8,10,11], orofacial [12] and eye movements in the horizontal or vertical axis [13]. Somatosensory modulation of tinnitus may be associated with underlying somatic complaints [8]. When tinnitus appears to be preceded or closely related to a somatic disorder, and therefore related to problems with the musculoskeletal system, and not the ear, the term “somatic tinnitus” or “somatosensory tinnitus” is used [14].

Animal studies have demonstrated that the etiology of somatic tinnitus involves cortical neuroplasticity, initiated by subcortical changes in brainstem nuclei receiving both somatosensory and auditory inputs [9]. In humans, somatic tinnitus is often a direct consequence of an injury or insult to the cervical spine (whiplash, concussion) or temporomandibular joint dysfunction [15]. The temporomandibular joint and cervical spine, together with the head, appear to be the musculoskeletal anatomical regions most commonly underlying somatic tinnitus, followed by eye movements and limbs [16]. This review aims to evaluate the prevalence of somatosensory modulation of the cervical spine and temporomandibular joint in subjects with tinnitus.

## 2. MATERIALS AND METHODS

### 2.1 Review protocol

The PRISMA Statement [17] systematic review reporting guidelines were used for this work.

### 2.2 Question of the review

Summarize the prevalence of somatosensory modulation of the cervical spine and temporomandibular joint in subjects with tinnitus.

### 2.3 Study eligibility criteria

Having identified the objective of the review, the following eligibility criteria were determined:

1. <u>Population</u>: adults with tinnitus.
2. <u>Studio design</u>: Observational studies were included to investigate the prevalence of somatosensory modulation of the cervical spine and temporomandibular joint in subjects with tinnitus.
3. <u>Tongue</u>: No restrictions have been applied.
4. <u>Age</u>: No restrictions have been applied.
5. <u>Time limit</u>: no time limit has been entered.

### 2.4 Analyzed databases

The databases investigated for the review were the following:

- Base
- Pubmed
- Scopus
- Web of Science

The search identified all articles indexed up to February 19, 2023. So this date is the upper limit of eligibility in this review.

### 2.5 Search strategies

To identify the keywords with which to construct the search string, the model with the acronym PECO was used, in which the patient or pathology (P), the exposure (E), the control with which to compare the exposure (C), and the outcome we are interested in (O). In the specific case of this review, the question did not include a control with which to compare the exposure factor; therefore, in this case, the CEEC has become PEO:

- P: subjects with tinnitus
- E: cervical spine and temporomandibular joint
- O: prevalence of somato-sensory modulation

The keywords have been chosen based on the review question posed, to obtain a search that is sensitive to the latter but not too specific to be able to sift through all the available literature.

### 2.6 Search string used

The search string was carefully designed to capture all potentially eligible records relating to the prevalence of somatosensory modulation of the cervical spine and temporomandibular joint in subjects with tinnitus. The terms were explored both as MeSH Terms (if they existed as such) and as free words and the Boolean operators OR, AND were used. The same string was used for all databases selected for this study (Table 1).

**Table 1.**
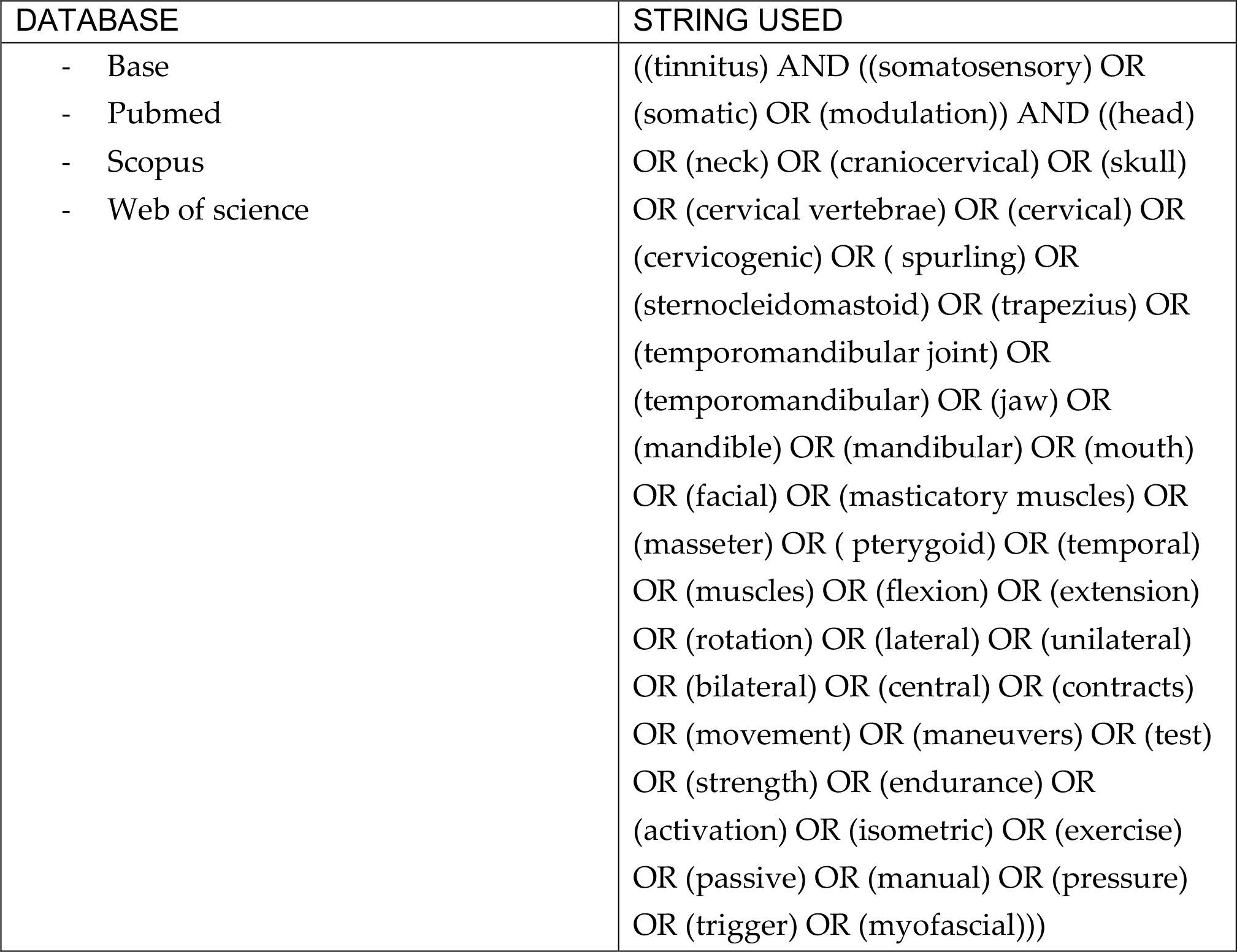
Databases analyzed and search string used.

| DATABASE | STRING USED |
| --- | --- |
| - Base<br>- Pubmed<br>- Scopus<br>- Web of science | ((tinnitus) AND ((somatosensory) OR (somatic) OR (modulation)) AND ((head) OR (neck) OR (craniocervical) OR (skull) OR (cervical vertebrae) OR (cervical) OR (cervicogenic) OR (spurling) OR (sternocleidomastoid) OR (trapezius) OR (temporomandibular joint) OR (temporomandibular) OR (jaw) OR (mandible) OR (mandibular) OR (mouth) OR (facial) OR (masticatory muscles) OR (masseter) OR (pterygoid) OR (temporal) OR (muscles) OR (flexion) OR (extension) OR (rotation) OR (lateral) OR (unilateral) OR (bilateral) OR (central) OR (contracts) OR (movement) OR (maneuvers) OR (test) OR (strength) OR (endurance) OR (activation) OR (isometric) OR (exercise) OR (passive) OR (manual) OR (pressure) OR (trigger) OR (myofascial))) |

### 2.7 Selection Process

Once the search was performed, a three-step process examined all records according to eligibility criteria: first by reading the title, second by reading the abstract, and third by reading the full text. The full text was read for all potentially relevant papers that appeared to meet the inclusion criteria or for which there was insufficient information in the title and abstract to make a firm decision. The study selection process was performed by a single author (MS) under the supervision of a second author (AG). At each stage any discrepancies were reviewed by a third author (PB) and a decision was made after discussion.

### 2.8 Data Collection Process

The data collection process was developed on 14 studies by two authors (MS-AG). The final data included were: year of study, author, population, inclusion and exclusion criteria, maneuvers, and results.

### 2.9 Summary of data

The primary purpose of this systematic review was to provide a summary of the percentage prevalence of somatosensory modulation of the cervical spine and temporomandibular joint in subjects with tinnitus.

## 3. RESULTS

### 3.1 Studies included

The identified strings produced a total (n) of 2442 articles divided as follows:

- 597 articles from Pubmed
- 638 articles from Scopus
- 518 articles from Web of Science
- 689 articles from Embase

First of all, duplicate articles were excluded (1301 articles), common to the searches carried out on the various databases, after which a selection was made by title from which 1070 articles were excluded that did not show relevance to the research question and/or did not comply with the eligibility criteria defined in paragraph 2.3.

Subsequently, from the 71 remaining, 53 articles were excluded after reading the abstract, because they were not relevant to the purpose of the review and/or to the eligibility criteria. A further selection was made below by analyzing the full text of the remaining articles, excluding 4 so that the articles compliant with the eligibility criteria, on which the review was built were 14:

1. Ralli M. et al (2017)[18]
2. Won JY. Et al (2013)[19]
3. Vielsmeier V. et al (2012)[20]
4. Simmons R. et al (2008)[13]
5. Ward J. et al (2015)[21]
6. Levine RA. Et al (2003)[22]
7. Ralli M. et al (2016)[23]
8. Bezerra R. et al (2008)[24]
9. Sanchez TG et al (2002)[14]
10. Sanchez TG et al (2007)[25]
11. Lee HY et al (2020)[26]
12. Pinchoff RJ et al (1998)[12]
13. Abel MD et al (2004)[27]
14. Theodoroff SM et al (2022)[28]

For a schematization of the study selection process and a description of the reasons why they were excluded, please refer to the Flow chart, shown in Figure 1.

**Figure 1.**
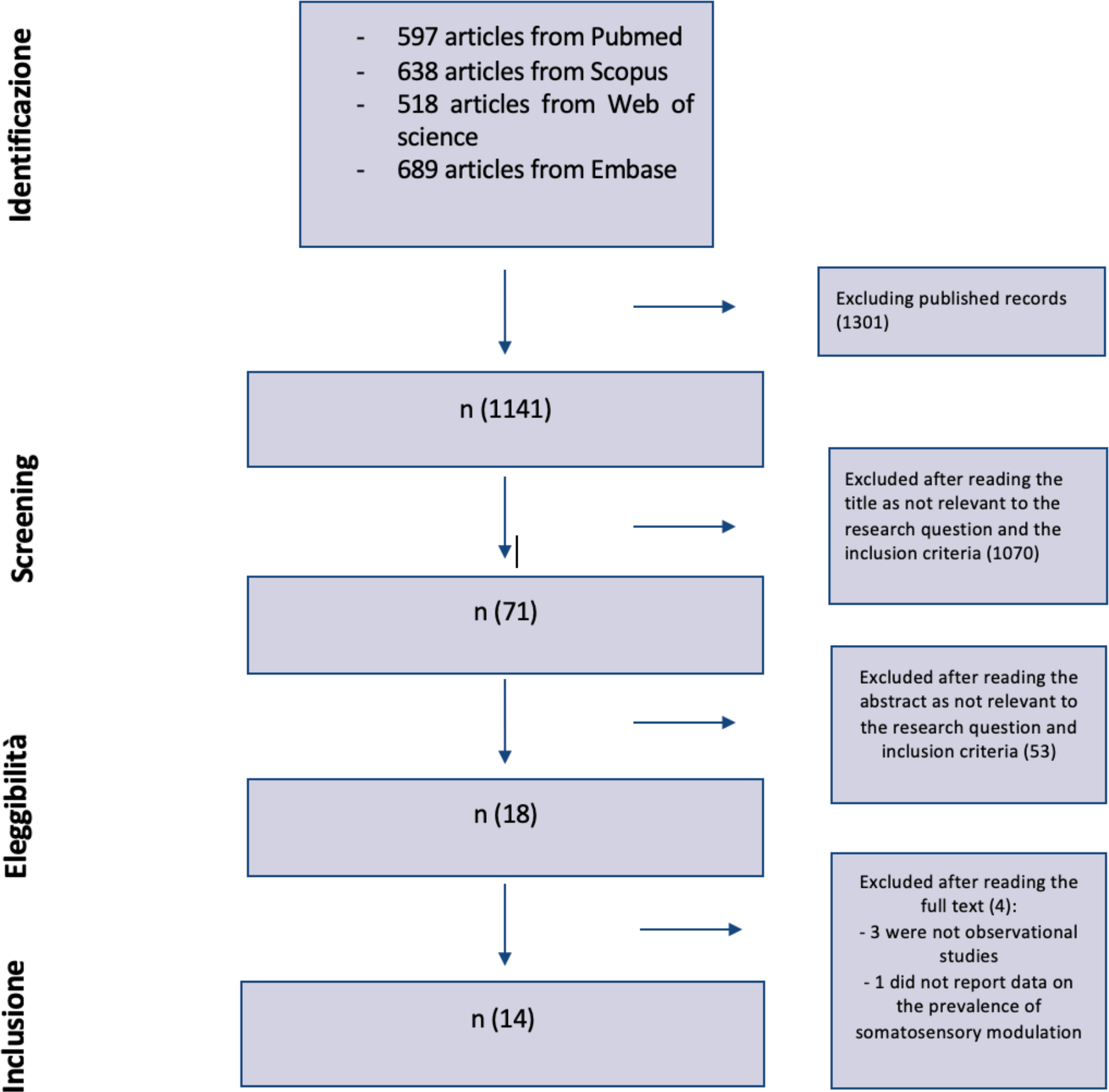
Flowchart.

### 3.2 Influence of TMJ and cervical spine in somatosensory modulation

The somatosensory modulation of tinnitus originates from the complex somatosensory-auditory interactions also deriving from anatomical regions such as the TMJ, cervical spine, head, eye or limb movement [8,10,11].

Five studies reported the prevalence of tinnitus modulation with one or more TMJ maneuvers, three studies with one or more cervical spine maneuvers, and seven studies reported combined movements involving both somatic regions mentioned above (see table 2).

**Table 2.** Prevalence of somatosensory modulation concerning the temporomandibular joint (TMJ) and cervical spine (CR).

| Author | Year | Population | Specific inclusion criteria | Somatic maneuvers | Somatic region | Prevalence of modulation(%) |
| --- | --- | --- | --- | --- | --- | --- |
| Pinchoff RJ et al | 1998 | 93<br>(Average age: 53 years) | -Somatic tinnitus (self-attributed diagnosis using questionnaires) | Do not specify (self-administered remotely) | ATM | 51 |
| Vielsmeier V. et al | 2012 | 261<br>(Average age: 50) | -Somatic tinnitus (medical diagnosis)<br>- ATM disorders (self-reported through questionnaires) | Do not specify (self-administered remotely) | ATM | 48 |
| Ralli M. et al | 2016 | 310<br>(Average age: 50) | -Somatic tinnitus (medical diagnosis)<br>-Normal hearing threshold (complete ENT exam)<br>- TMJ and/or RC disorders (medical evaluation) | ATM (5 manoeuvres)<br>- Resisted tests performed in person by the patient<br>RC (14 maneuvers)<br>- Active movements performed in person by the patient (4 maneuvers)<br>-Resisted tests performed by the examiner (10 manoeuvres) | ATM<br>RC<br>ATM, RC | 26.77<br>14.19<br>38.71 |
| Ralli M. et al | 2017 | 172<br><u>82 patients</u>with somatic tinnitus+ Hyperacusis (ASI), average age 43 years<br><u>90 patients</u>with somatic tinnitus without hyperacusis (AS), average age: 39 years | -Somatic tinnitus (medical diagnosis)<br>-Hyperacusis (complete ENT exam) | ATM (5 manoeuvres)<br>- Resisted tests performed in person by the patient<br>RC (14 maneuvers)<br>- Active movements performed in person by the patient (4 maneuvers)<br>-Resisted tests performed by the examiner (10 manoeuvres) | ATM<br>RC<br>ATM, RC | ASI: 29.54<br>AS: 39.40<br>ASI: 11.36<br>AS: 33.33<br>ASI: 59.9<br>AS: 27.27 |
| **Theodoroff SM et al** | 2022 | 205<br>(Average age: unspecified) | -Veterans with somatic tinnitus (self-assigned diagnosis using questionnaires) | ATM (7 manoeuvres)<br>-Resisted tests and active movements performed by the patient at a distance<br>RC (4 maneuvers)<br>-Resisted tests and active movements performed by the patient at a distance | ATM<br>RC | 87.80<br>77.07 |
| **Won JY. et al** | 2013 | 163<br>(Average age: 56 years) | - Somatic tinnitus (medical diagnosis) | ATM (9 manoeuvres)<br>-Resisted tests performed by the examiner<br>RC (10 maneuvers)<br>-Resisted tests performed by the examiner | ATM, RC | 57.1 |
| **Bezerra Rocha et al** | 2008 | 68<br>(Average age: 53 years) | - Constant tinnitus (unilateral or bilateral) for at least 3 months (medical diagnosis) | -Pressures by the examiner (9 muscles analysed) | ATM, RC | 55.9 |
| **Sanchez TG et al** | 2002 | 121<br>(Average age 52 years) | -Somatic tinnitus (medical diagnosis) | ATM (1 manoeuvre)<br>-Resisted test performed in person by the patient<br>RC (8 self-administered manoeuvres)<br>- Resisted tests performed in person by the patient | ATM, RC | 61.2 |
| **Sanchez TG et al** | 2007 | 38<br>(Average age 56 years) | - Unilateral or bilateral tinnitus (medical diagnosis) | ATM (1 manoeuvre)<br>-Resisted test performed in person by the patient<br>RC (8 maneuvers)<br>- Resisted tests performed in person by the patient | ATM, RC | 57.9 |
| Lee HY et al | 2020 | 81<br>(Average<br>age: 52 years) | -Subjective non-<br>pulsatile tinnitus<br>lasting >3 months<br>(medical diagnosis) | ATM (9<br>manoeuvres)<br>-Resisted tests<br>performed by the<br>examiner<br>RC (10 maneuvers)<br>-Resisted tests<br>performed by the<br>examiner | ATM, RC | 61.7 |

As far as modulation following TMJ maneuvers is concerned, the study by Pinchoff RJ et al of 1998 [12] reports a prevalence of 51% while the study by Vielsmeier et al of 2012 [20] of 48% on 261 patients.

The study by Ralli M et al in 2016 [23] took into consideration 310 patients of which 302 with a positive history of TMJ and/or cervical spine disorders. Tinnitus volume could be modulated in 79.67% of patients. Of these, in 26.77% of cases, the modulation occurred with one or more maneuvers of the ATM, in 14.19% with one or more maneuvers of the cervical spine, and in 38.71% using maneuvers involving both districts.

Ralli M. et al in a 2017 study [18] evaluated the prevalence of modulation in patients with somatic tinnitus with hyperacusis (82) and without hyperacusis (90).

In the tinnitus and hyperacusis group, 29.54% were able to modulate tinnitus by performing one or more TMJ maneuvers, 11.36% by one or more cervical spine maneuvers, and 59.09% by one or more plus maneuvers involving both districts.

In the group with somatic tinnitus without hyperacusis, 39.40% managed to modulate tinnitus with one or more TMJ maneuvers, 33.33% with one or more cervical spine maneuvers, and 27.27% with one or more plus maneuvers involving both districts.

The 2022 study by Theodoroff SM et al [28] reports data obtained through telephone screening among veterans with tinnitus in which somatic tinnitus was found in 205 of 292 patients who were able to perform somatic maneuvers at home. Of these 205, 158 (77.07%) had a change with neck maneuvers and 180 (87.80%) with TMJ maneuvers.

Unlike the previous ones, the following studies report unified data regarding the prevalence of tinnitus modulation obtained with maneuvers of the cervical spine and TMJ.

In the study by Won JY et al from 2013 [19] the results are presented in terms of the total number of ears with tinnitus and no patients as somatic stimuli usually modulate tinnitus in the ipsilateral ear rather than both ears at the same time. A total of 217 ears were studied and tinnitus was found to be modulated by at least one maneuver out of 124 ears (57.1%). In Sanchez TG’s 2002 study [14] tinnitus modulation induced by maneuvers involving the TMJ and cervical spine muscles occurred in 74/121 (61.2%) patients; while, in a subsequent 2007 study [25], among the 38 patients studied, 22 (57.9%) experienced tinnitus modulation in the initial test (T1) during at least one TMJ muscle contraction maneuver and/ or of the neck.

Lee HY et al 2020 study [26] reports that 51/81 patients (61.7%) modulated their tinnitus using one or more neck and/or TMJ maneuvers.

Among the studies taken into consideration was also included the study by Bezerra Rocha et al, 2008 [24] which differs from the others in that it concerns the modulation of tinnitus with pressure on myofascial trigger points; in it a prevalence of modulation of 55.9% was found on 68 participants.

## 4. DISCUSSIONS

Somatic modulation is a common condition in patients with tinnitus. Levine (1999) [8] has defined it as a ‘‘functional attribute’’ of tinnitus and several authors have found a large capacity for somatic modulation of tinnitus in different series of patients, ranging from 65.3 to ‘83.3% [13,14,19,22,25].

As regards the modulation specifically attributable to TMJ manoeuvres, in the studies by Pinchoff RJ et al [12] and by Vielsmeier V. et al [20] an average prevalence of 49.5% is obtained, which would seem clearly optimistic compared to what was reported in the study by Ralli M. et al in 2016 [23], according to which the prevalence of modulation, taking into account only the maneuvers concerning the TMJ, stands at just over half (26%). Even more optimistic would seem the data obtained by Theodorff SM et al [28] with a modulation percentage reaching almost 90%. However, the methods of execution of the studies must be taken into consideration, as in those of Pinchoff RJ et al, of Vielsmeier V. et al and in that of Theodoroff SM et al the patient found himself performing the different somatic maneuvers without the supervision of an examiner, unlike what happened in the studies by Ralli M. et al of 2016 and 2017, where the different maneuvers used were performed in part under the supervision supervision by the examiner and partly by the examiner. Compared to its previous 2016 study, the 2017 study [18] shows a higher mean TMJ prevalence (34%), with however a significant difference found between patients with and without hyperacusis; the latter would seem to have a greater probability of modulating tinnitus. where the different maneuvers used were performed partly under the supervision of the examiner and partly by the examiner. Compared to its previous 2016 study, the 2017 study [18] shows a higher mean TMJ prevalence (34%), with however a significant difference found between patients with and without hyperacusis; the latter would seem to have a greater probability of modulating tinnitus. where the different maneuvers used were performed partly under the supervision of the examiner and partly by the examiner. Compared to its previous 2016 study, the 2017 study [18] shows a higher mean TMJ prevalence (34%), with however a significant difference found between patients with and without hyperacusis; the latter would seem to have a greater probability of modulating tinnitus.

Instead, as regards the studies investigating the modulation of the cervical spine, we have extremely variable data: in the study by Ralli M. et al of 2016 we have a percentage of 14%, in the study by Theodoroff SM et al a percentage of 77%, while intermediate data can be found in the study by Ralli M. et al of 2017. Also in this case a significant difference is observed between patients with and without hyperacusis who show modulation percentages of 11% and 33% respectively.

It can be seen how Theodoroff SM’s datum is always very different from the others, showing clearly higher percentages. These results could be influenced by specific factors of the analyzed population, as well as by the execution modalities of the maneuvers as previously observed.

In the five studies that take into consideration the prevalence of modulation with the combination of maneuvers of the TMJ and the cervical spine without providing separate data, very similar percentages are found: in two studies around 57% (Sanchez TG et al of 2007 [25], Won JY et al [19]), in two others of about 61% (Sanchez TG et al of 2002 [14], Lee HY et al [26]) and in the last one of 55.9% (Bezerra R. et al [24]). It should be noted that the populations analyzed in the pairs of studies with similar percentages of modulation have the same mean age, which corresponds to 52 years for the studies by Sanchez TG et al of 2002 and Lee HY et al, and to 56 years for the studies by Sanchez TG et al of 2007 and Won JY et al. It could be a coincidence or younger patients have a better chance of modulating tinnitus. In favor of this could be the fact that this difference is not attributable, for example, to the methods of administration of the manoeuvres, since in both pairs of studies there is a study in which the maneuvers are performed by the examiner and a study in which which are performed by the patient under the supervision of the examiner and, despite this, the prevalence of the modulation stands at the same values.

Overall, the 5 studies cited above report on average a higher prevalence of modulation following combined TMJ and cervical spine maneuvers than the studies by Ralli M. et al. In his 2016 and 2017 studies, there is a prevalence of approximately 39% and 44% respectively, in the latter case as a unified figure between patients with and without hyperacusis. Here too there is a significant difference between patients with and without hyperacusis, who obtained modulation respectively in 59.9% and 27.27% of the cases, so in this case, unlike what was found by taking consider individual districts separately, those with hyperacusis respond more to somatic maneuvers.

A part of the studies included in this review does not report the exclusive data only for the TMJ and cervical spine but rather the percentage of general modulation for the different somatic regions analyzed in the different studies (Table 3).

**Table 3.** General prevalence of somatosensory modulation for the different somatic regions analyzed in the different studies.

| Author | Year | Population | Specific inclusion criteria | Somatic maneuvers | Somatic region | Prevalence of modulation (%) |
| --- | --- | --- | --- | --- | --- | --- |
| Simmons R. et al | 2008 | 45<br>(Average age: 65.2 years) | - Somatic tinnitus (self-attributed diagnosis through questionnaires) | 42 unspecified maneuvers (active and resisted movements performed in person by the patient and passive movements performed by the examiner) | ATM, head, RC | 78 |
| Ward J. Et al | 2015 | 671<br>(Average age: 56.3 years) | - Tinnitus (self-attributed diagnosis through questionnaires) | Unspecified (remotely self-administered) | ATM, head, RC | 16.1 |
| Abel MD et al | 2004 | 60<br>(Average age: 52 years) | - Tinnitus (medical diagnosis) | ATM (10 manoeuvres)<br>- Active and resisted movements performed in person by the patient<br>RC (10 maneuvers)<br>- Resisted tests performed in person by the patient<br>Limbs (5 maneuvers)<br>- Resisted tests performed in person by the patient | TMJ, head, RC, limbs | 83.3 |
| **Levine RA. et al** | 2003 | 75<br>(Average age: 46 years) | - Tinnitus (self-diagnosed) | ATM (10 manoeuvres)<br>-Active and resisted movements performed in person by the patient<br>RC, head (10 maneuvers)<br>- Resisted tests performed in person by the patient<br>Limbs (5 maneuvers)<br>- Resisted tests performed in person by the patient | TMJ, head, RC, limbs | 79 |

The four studies consider the modulation relating to movements that have affected several districts at the same time. The districts taken into consideration are the TMJ, head and cervical spine in the studies of Simmons R. et al [13] and Ward et al [21], while in those of Abel MD et al [27] and Levine RA et al [22] additional limb movements were performed.

The prevalences found were in general quite high (78% [13],83.3% [27],79% [22]), except for the study by Ward et al, where on the contrary we have a prevalence of 16, 1%. It is important to underline that this is also the only study that reports a percentage obtained through remote self-administered maneuvers.

A final consideration can be made regarding the studies by Abel MD et al and Levine RA et al in which the addition of limb movements changed the percentage slightly compared to the study by Simmons R. et al, since it suggests a role less relevant than this district in the somatic modulation of tinnitus.

## 5. CONCLUSIONS

The present study confirmed that tinnitus perception and intensity can be somatically modulated in a subpopulation of individuals. The 14 studies included in the review show great heterogeneity from the point of view of the populations and districts analyzed, which made it difficult to understand the different relationships between tinnitus and the specific somatic regions under study. However, the cervical spine and the TMJ play a key role in the modulation of tinnitus and, based on the data collected on individual districts, the TMJ appears to be the somatic region that is most capable of modulating tinnitus. However, further studies are needed to investigate somatosensory modulation in individual districts and to include more specific inclusion criteria regarding age, comorbidities, and other factors that can significantly affect the results, to obtain more reliable data and exhaustive.

## Data Availability

All data produced in the present study are available upon reasonable request to the authors

